# Ayahuasca-Inspired DMT/HAR Formulation Reduces Brain Differentiation Between Self and Other Faces

**DOI:** 10.1101/2024.10.28.24316308

**Authors:** Dila Suay, Helena D. Aicher, Micheal Kometer, Michael J. Mueller, Luzia Caflisch, Alexandra Hempe, Camilla P. Steinhart, Claudius Elsner, Ilhui A. Wicki, Jovin Müller, Daniel Meling, Dario A. Dornbierer, Milan Scheidegger, Davide Bottari

**Author notes:** **Corresponding author:** Dila Suay, IMT School for Advanced Studies, Lucca, Italy, Piazza San Francesco 19, 55100, Lucca, Italy. Equal Contribution (Milan Scheidegger, Davide Bottari).

## Abstract

**Background:** Psychedelics are known to profoundly alter perception and self-referential processing, yet their specific effects on face recognition—a key aspect of social cognition—remain underexplored.

**Objective:** This study investigates the effects of an ayahuasca-inspired novel DMT/HAR (N,N-dimethyltryptamine/Harmine) formulation and Harmine alone on face recognition and self-referential processing, as measured by event-related potentials (ERPs).

**Methods:** In a within-subject, double-blind, placebo-controlled design, 31 healthy male participants underwent EEG recording during a visual oddball task involving Self, Familiar, and Unknown Faces. The study compared the effects of a DMT/HAR formulation, harmine alone, and placebo on key visual ERP components: P1, N170, and P300.

**Results:** DMT/HAR increased P1 amplitude and decreased N170 amplitude across all face categories, indicating enhanced early visual processing and disrupted face structural encoding. DMT/HAR also reduced P300 amplitude specifically for self-faces, diminishing neural differentiation between self and other faces.

**Conclusion:** The DMT/HAR formulation significantly blurs the neural distinction between self and other faces, suggesting a potential mechanism by which psychedelics diminish attentional focus on self-referential information, thereby enhancing empathy and unity. These insights into serotonergic modulation of face recognition could inform therapeutic strategies for disorders characterized by altered self-processing.

## 1. Introduction

Psychedelics (serotonergic hallucinogens) temporarily alter perception and impact a multitude of cognitive processes, including changes in consciousness and the subjective perception of oneself. One of the most striking characteristics of the psychedelic experience is the alteration of sensory processing, particularly visual perception (Kometer & Vollenweider, 2016). These alterations are not only central to the psychedelic experience but also serve as a crucial window into understanding how psychedelics modulate the brain’s visual processing systems. Psychedelics such as psilocybin, lysergic acid diethylamide (LSD), and N, N-dimethyltryptamine (DMT) exert their hallucinogenic effects primarily through agonism of serotonin (5-HT) receptors, which are densely distributed in cortical pyramidal neurons and key visual processing areas (López-Giménez & González-Maeso, 2018). The intricate role of these receptors in visual perception is further underscored by the fact that antagonists like ketanserin can block the perceptual effects of these psychedelics, thereby validating the importance of 5-HT2 receptor activation in generating visual hallucinations (Quednow et al., 2012). The impact of psychedelics on visual perception is evident at multiple levels of brain function. Previous neuroimaging studies showed that psychedelics alter visual processes at several levels, such as augmenting functional connectivity between the primary visual cortex and the rest of the brain (Carhart-Harris et al., 2016), boosting fMRI BOLD signals in visual areas at rest (de Araujo et al., 2012; Roseman et al., 2016), increasing bottom-up traveling waves (Alamia et al., 2020), and altering oscillatory activity (Kometer et al., 2013a; Muthukumaraswamy et al., 2013; Timmermann et al., 2019).

In parallel with broader functional changes, psychedelics also influence visual event-related potentials (ERPs), the P1 and N170 components of visual ERPs, revealing distinct effects on sensory and cognitive processing. The P1 component, occurring 50–100 milliseconds post-stimulus, exhibits increased amplitude under the influence of psychedelics, indicating enhanced early sensory processing potentially contributing to the vivid and intensified visual experiences reported by users (Kometer et al., 2011, 2013). Conversely, the N170 component, which is critical for the processing of complex visual stimuli like faces and objects, is disrupted by psychedelics (Schmidt et al., 2012; Kometer et al., 2011). This disruption, evident in altered N170 amplitude and latency, correlates with the visual distortions and hallucinations experienced during a psychedelic experience, reflecting how these substances interfere with the brain’s ability to integrate and interpret visual information (Kometer et al., 2011).

In addition to their effects on visual perception, psychedelics induce transient and dose-dependent reductions in one’s subjective experience of self, also known as ego dissolution (Letheby & Gerrans, 2017; Stoliker et al., 2022). During ego dissolution, the boundaries between subjective and objective realities blur, leading to a profound sense of oneness. This experience may offer therapeutic benefits, including improved mood, enhanced prosocial behavior, increased altruism, and overall well-being (Griffiths et al., 2008; Kometer et al., 2015). Research has consistently demonstrated that psychedelics such as psilocybin, LSD, and DMT disrupt the default mode network (DMN) – a network of brain regions, including the posterior cingulate cortex, medial prefrontal cortex (mPFC), and inferior parietal lobule, which is crucial for self-referential processing (Qin & Northoff, 2011). Studies investigating the link between 5-HT2A receptor activation and ego dissolution have found that increased ego dissolution correlates with reduced DMN integrity (Carhart-Harris et al., 2014, 2016; Lebedev et al., 2015; Palhano-Fontes et al., 2015). Similarly, changes in the P300 ERP component – associated with cognitive resource allocation and self-referential processing – correlate with enhanced feelings of connectedness and altered perceptions of meaning during psychedelic experiences (Bravermanová et al., 2018; Smigielski et al., 2020).

Building on the observed psychedelic-induced alterations in visual and self-referential processing, this study aims to extend these findings to the domain of face recognition – a field that has yet to be thoroughly explored in the context of psychedelics. To address this gap, the present study was designed to explore the effects of an ayahuasca-inspired formulation combining DMT and harmine on face recognition. Ayahuasca is a traditional South American plant brew known for its potent psychoactive properties. This brew typically contains two key ingredients: beta-carboline alkaloids such as harmine or harmaline derived from the *Banisteriopsis caapi* vine and the hallucinogenic compound DMT sourced from the *Psychotria viridis* plant (McKenna et al., 1984). When ingested orally, DMT alone is rendered inactive due to rapid first-pass metabolism by monoamine oxidase A (MAO-A) in the body. To counteract this, ayahuasca includes beta-carbolines that inhibit MAO-A, thereby preventing the breakdown of DMT and allowing it to produce its psychoactive effects (McKenna, 2004). In this study, we employed a novel formulation that combines buccally administered harmine with intranasally administered DMT (Dornbierer et al., 2023) to evaluate its impact on face recognition using a within-subject, double-blind, placebo-controlled design with three randomized conditions. During each session, participants’ electrophysiological activity (EEG) was recorded while they performed a visual oddball task with stimuli categorized as Self, Familiar, and Unknown Faces. We investigated the effects of the DMT/HAR formulation and harmine alone compared to placebo on key visual event-related potentials, including P1, N170, and P300. Additionally, we explored the relationship between these electrophysiological measures and critical features of the psychedelic experience, such as elementary and complex imagery scores. We hypothesized that DMT/HAR, but not harmine, would significantly blur the neural boundaries between self and other faces, potentially providing new insights into the neural underpinnings of face recognition under psychedelic influence.

## 2. Methods

### 2.1. Experimental design

This study employed a within-subject, double-blind, placebo-controlled design with three sessions of counterbalanced drug administrations:DMT/HAR, harmine alone (HAR), placebo (PLA), each separated by a two-week washout period. This experiment is part of a larger series of studies, detailed in Aicher & Mueller et al. (2024) and was conducted at the peak drug effect, 60 minutes post initial DMT (or placebo) administration. The experimental task lasted for 30 minutes, while the entire duration of study was 360 minutes.

### 2.2. Participants

Thirty-six right-handed healthy male volunteers (20–40y, BMI 18.5–30) were recruited for this study. Inclusion criteria comprised: (a) (a) no current or previous history of neurological disorders or psychotic or bipolar disorders, and no family history of psychotic or bipolar disorders (b) fluent German language normal or corrected-to-normal vision (d) normal hearing (e) little to no experience with hallucinogenic drugs (max. 15 lifetime). All participants underwent medical and psychological screening upon completing the initial telephone screening. On-site screenings were conducted in the same room as the experiments, ensuring familiarity with the surroundings, which was found conducive to the psychedelic experience (Hartogsohn, 2017). During the screenings, participants completed a training session to familiarize themselves with the tasks. The study enrolled 36 participants, Five participants voluntarily withdrew from the study, and one was excluded due to technical difficulties during data collection. Consequently, the final sample analyzed for this article comprised 30 participants. The study was conducted according to the World Medical Association Declaration of Helsinki. The study was approved by the Cantonal Ethics Committee of the Canton of Zurich (Basec-Nr. 2018-01385) and Swiss Federal Office of Public Health (BAG-Nr. (AB)-8/5-BetmG-2019/008014), and it was registered on clinicaltrials.gov (ID: NCT04716335). Participants gave written informed consent and were reimbursed commensurate with the time invested for participation (320.-CHF total, or 60.-per completed intervention day).

### 2.3. Drug and dosing

A counterbalanced order of (i) DMT/HAR, (ii) harmine alone (HAR), and (iii) placebo (PLA) were administered on three separate test days with identical procedure with a minimum 2 weeks of interval to prevent carry-over effects. (i) For the DMT/HAR condition, we used a nasal spray for standardized administration of DMT (isolated from *Mimosa tenuiflora*) and a buccal orodispersible tablet containing the monoamine oxidase inhibitor harmine (purchased from Sigma-Aldrich / Merck KGaA) (Dornbierer et al., 2023). Thirty minutes after buccal administration of 100 mg harmine, the DMT nasal spray was administered at intervals of 15 minutes. Each administration contained 10 mg of DMT, and the EEG experiment started 60 minutes after the first DMT administration when participants reached a total dose of 50 mg. This repeated-intermittent dosing regimen demonstrated that participants exhibited good self-control and coped well with the drug experience in an experimental setting (Aicher & Mueller et al., 2024). (ii) For the harmine condition (HAR), participants were administered 100 mg harmine buccally, along with a placebo nasal spray which was indistinguishable from drug with respect to taste, smell, and appearance. (iii) In the placebo condition (PLA), both orodispersible tablet and nasal spray were without active molecules and mimicked the natural drugs’ appearance and taste.

### 2.4. Procedure

#### 2.4.1. Measures of Acute Subjective Effects

Participants’ subjective states of consciousness were evaluated using the 5-Dimensions Altered State of Consciousness questionnaire (5D-ASC) (Dittrich, 1998). The 5D-ASC is a well-validated self-rating visual analogue scale (VAS) developed to quantify altered states of consciousness and has been widely employed to assess the structure and phenomenology of psychedelic experiences (e.g., Studerus et al., 2010; Kometer et al., 2015). Before the experiment, participants completed VAS ratings using a digital display screen and button press format. This study specifically utilized the elementary imagery and complex imagery scores from the 11 sub-dimensions of the 5D-ASC to focus on visual perception (Studerus et al., 2010). These items included specific questions related to the intensity and nature of visual experiences, such as “How intense were your visual experiences?” and “Did you experience complex visual imagery?” This approach allowed for real-time assessment of participants’ visual perception during the experiment.

#### 2.4.2. Stimuli

In the EEG experiment, the set of face stimuli consisted of 396 images pertaining to four different categories: (i) self faces, (ii) familiar faces, (iii) unknown faces (n=120 in each condition), and (iiii) target (n=36). (i) For self-face images, participants were photographed during on-site medical screening (Sony DSLR-A350). (ii) Unknown-face images were selected from the Chicago Face Database (Ma et al., 2015). (iii) For the familiar-face images, we chose to use celebrity pictures instead of participants’ own family or friends to avoid additional biases that can arise in self-referential information processing from ingroup membership (Platek & Kemp, 2009). A total of 262 male celebrity pictures were collected over the internet, including the faces of politicians, musicians, athletes, and actors. During the on-site medical screening, participants went through a pre-experiment familiarity test, where they rated each celebrity face as (a) I do not recognize, (b) I recognize somehow, (c) I recognize, and (d) I can name the person. Based on the ratings, 120 celebrity pictures were selected from the (c) and (d) categories to create participant-specific familiar-face stimuli. (iiii) Finally, scrambled images of participants’ self-faces were used as a target (all image pixels were randomly oriented and placed).

All face stimuli were presented in a frontal view with a neutral expression and converted to greyscale. An ellipse mask (562 × 762 pixels resolution, 600 DPI) was applied to each face, cropping the exterior features such as hairline and ears, leaving only the inner facial features against a black background. This was achieved using the GNU Image Manipulation Program (GIMP) (GNU Image Manipulation Program, 1995). To maintain consistency, the contrast and luminance histograms of the images were standardized using the default function provided in the SHINE toolbox (Willenbockel et al., 2010), which was implemented within the MATLAB R2020a environment (The MathWorks, Inc., 2020). The spatial frequencies of the images across different face categories—namely, Self (Mean = 0.179, SD = 0.079), Familiar (Mean = 0.169, SD = 0.042), and Unknown (Mean = 0.175, SD = 0.044)—were analyzed. Statistical analysis using ANOVA revealed no significant differences in spatial frequencies between these face categories (ANOVA: *t*(2,419) = 1.245, *p* = 0.289). Stimuli were electronically presented using E-Prime 3.0 software (Psychology Software Tools, Inc., 2016) at a fixed viewing distance of 90 cm on a 25-inch screen. This setup provided a visual angle of 11° x 15° (horizontal x vertical). Responses were collected via the E-Prime Chronos Device.

#### 2.4.3. Experiment Procedure

Subjects were instructed to keep their gaze on the fixation cross placed at the center of the screen for the entire block duration and were asked to press a button with their right index finger when the scrambled face (target) appeared. Targets could occur with a probability between 8 and 12%. This procedure ensured that participants attended each visual image and prevented eliciting motor responses for the relevant face categories (self-faces, familiar faces, unknown faces). The stimulus duration was 100 ms, and the interstimulus interval varied randomly between 1 and 1.4 seconds. The experiment was divided into three blocks, interleaved by a 3-minute break, and the overall duration was 30 minutes. To prevent habituation, stimuli were presented pseudo-randomly so that no more than three faces of the same category could consecutively occur. On average, participants performed target detection with 98% accuracy. All conditions were presented in each experimental session (drug condition): DMT/HAR, HAR, and PLA. From this point forward, we will use the following abbreviations: SELF for Self, FAM for Familiar, and UNK for Unknown. Therefore, the condition labels are as follows: DMT/HAR _SELF_, HAR_SELF_, PLA_SELF_, DMT/HAR _FAM_, HAR_FAM_, PLA_FAM_, DMT/HAR _UNK_, HAR_UNK_, PLA_UNK._

#### 2.4.4. EEG recordings

Continuous EEG was recorded with the BioSemi Active 2 system with 64 Ag-AgCl pin-type active electrodes mounted on an elastic cap, according to the international 10-20 system. Eight additional flat-type active electrodes were used for body surface measurements. For horizontal and vertical electrooculography (EOG), electrodes were placed on each eye’s outer canthus and the left eye’s supraorbital and infraorbital. For electrocardiography (ECG), electrodes were placed on the end of the left collarbone and the lower left rib. Additional electrodes were placed on the right and left mastoid bones. Data were referenced to the Common Mode Sense (CMS) and the Driven Right Leg (DRL) electrodes, while the average voltage was kept in the range of ±40 mV signal using Biosemi’s electrodes offset tool. Signals were recorded with a 0.1 Hz high-pass filter and a 100 Hz low-pass filter, with a sampling rate of 2048 Hz. Triggers were sent to the BioSemi software and recorded using a parallel port along with the EEG data.

#### 2.4.5. EEG preprocessing

The preprocessing was performed offline using EEGLAB (Delorme & Makeig, 2004) within the MATLAB environment (The Math Works, Inc., 2020). Data were preprocessed with a semi-automatic pipeline implemented in MATLAB (Stropahl et al., 2018). Data were re-referenced to the Fz electrode after they were imported. Stereotypical artifacts (eye blinks, lateral eye movements, and heartbeats) were detected using an infomax independent component analysis (ICA) (Bell & Sejnowski, 1995). To improve ICA decomposition, data were low-pass filtered (windowed sinc FIR filter, cut-off frequency 40Hz, filter order 256), resampled to 256 Hz and high-pass filtered (windowed sinc FIR filter, cut-off frequency 1Hz, filter order 512) and finally, segmented into consecutive dummy epochs of 1 second. The epochs with joint probabilities greater than 4 standard deviations were rejected (Bottari et al., 2020). ICA weights were then attributed to the unfiltered raw data (Strophal et al., 2015). Topographies of each component were plotted and visually inspected for artifacts (eye blinks, saccades, and heartbeat). Across all datasets, components representing the best stereotypical artifacts were used as a template for a semi-automated CORRMAP algorithm to identify ICA components with similar topography across all datasets (Viola et al., 2009). A correlation of the ICA inverse weights was computed, and the components with a correlation coefficient higher or equal to 0.8 were removed (from the raw data). Artifact-free data were low-pass filtered (windowed sinc FIR filter, cut-off frequency 40Hz, filter order 256), resampled to 256 Hz and high-pass filtered (windowed sinc FIR filter, cut-off frequency 0.1Hz, filter order 5120). Bad channels were interpolated with the spheric spline. The proportion of interpolated electrodes was kept at <10% for each participant. Data were re-referenced to the average. Data were segmented as a function of stimulus category (SELF, FAM, UNK) and conditions (DMT/HAR, HAR, PLA), into epochs containing 200 ms pre-stimulus and 800 ms post-stimulus. Within the same stimulus category/condition, epochs that had a higher joint probability of 4 standard deviations were excluded from further analysis, to remove epochs contaminated by non-stereotypical artifacts. Finally, epochs exceeding +-100 μV were also excluded. The number of removed epochs did not differ across conditions/stimulus categories (repeated-measures ANOVA (F (4, 116) = 0.35, p = 0.843, ηp^2^= 0.013; the mean and the ± SD of removed epochs for each conditions across stimulus categories were as follows: 5.9% ± 2.6 for DMT/HAR_SELF_, 5.8% ± 2.3 for DMT/HAR_FAM_, 5.7% ± 2.7 for DMT/HAR_UNK_; 5.4% ± 1.7 for HAR_SELF_, 5.4% ± 1.5 for HAR_FAM_, 5.5% ± 1.6 for HAR_UNK_; 5.8% ± 1.7 for PLA_SELF_, 5.6% ± 1.3 for PLA_FAM_, 5.4% ± 1.3 for PLA_UNK)_. Finally, epochs were baseline-corrected from -200 to 0 ms.

### 2.5. Statistical Approach

#### 2.5.1. Measures of Acute Subjective Effects

Elementary and complex imagery scores (5D-ASC) were analyzed using repeated measures analyses of variance (rmANOVAs; JASP) with the within-participant conditions as factors (DMT/HAR, HAR, PLA). The assumption of sphericity was tested using Mauchly’s test. If epsilon (ε) values were <0.75, Greenhouse-Geisser correction were used, and if they were >0.75, Huynh-Feldt correction was applied. When the F-statistics were significant, post-hoc paired t-tests were performed using a Bonferroni correction (adjusted alpha=0.017, to account for three tests).

#### 2.5.2. EEG experiment

First, we assessed the main effects of the drug conditions (DMT/HAR, HAR, PLA) on each face category (Self, Familiar, Unknown). We employed cluster-based nonparametric permutation F-statistics (MANOVA, Monte Carlo sampling method, 1000 iterations, cluster alpha *p* <0.05, maxsum criterion, cluster-based method for multiple comparison correction, minimum spatial extent 2 adjacent channels (Maris & Oostenveld, 2007) to determine whether the response to each face category differed across conditions. In case of significant effects in the MANOVA, we then employed cluster-based nonparametric permutation t-statistics (Monte Carlo sampling method, 1.000 iterations, two-tailed, cluster alpha p <0.025 (corresponding to a critical alpha level of 0.05 for two-tailed testing, accounting for both positive and negative clusters), maxsum criterion, minimum spatial extent 2 adjacent channels) between pairs of conditions. For both F- and t-statistics, analyses were run across all electrodes and within the whole-time window (0–600 ms). This data-driven approach allowed for identifying the significant difference between conditions without bias from a priori assumptions of a specific region of interest (ROIs) or time intervals while solving multiple comparison problems (Maris & Oostenveld, 2007). If the cluster-based p-value was less than 0.025, we rejected the null hypothesis that there was no difference between conditions.

Secondly, we examined the specific hypothesis that the modulation induced by DMT/HAR was greater for the self-face compared to both familiar and unknown faces. To this end, the interaction effect of drug conditions (DMT/HAR, HAR, PLA) and face stimuli (Self, Familiar, Unknown) was investigated by running a series of nonparametric cluster-based permutation t-statistics (Monte Carlo sampling method, 1.000 iterations, one-tailed for negative clusters, cluster alpha p <0.05, maxsum criterion, minimum spatial extent 2 adjacent channels) of the differential waves calculated by subtractions across face categories (e.g. DMT/HAR_SELF-FAM_ vs. PLA_SELF-FAM_ or DMT/HAR_SELF-UNK_ vs. PLA_SELF-UNK_), within the whole time-window (0–600 ms) across all electrodes.

#### 2.5.2. Correlations between EEG and Behavioral Data

To investigate the association between EEG and behavioral data, the Pearson correlation coefficients between EEG data and elementary/complex imagery scores were calculated using nonparametric cluster-based permutation t-statistics (ft_statfun_correlationT, Monte Carlo sampling method, 1.000 iterations, cluster alpha p <0.05, maxsum criterion, cluster-based method for multiple comparison correction, minimum spatial extent 2 adjacent channels) across all electrodes and time points (0–600 ms) (Oostenveld et al., 2011). Additionally, we analyzed the correlation between the behavioral data and P1, N170 and P300 ERP components with a hypothesis-driven approach using the same nonparametric cluster-based permutation t-statistics. Occipital electrodes (OZ, O1, O2) were chosen within 90 to 150 ms for P1, posterior electrodes (P8, PO8) were chosen within 160 to 190 ms for N170, and central-posterior electrodes (POZ, Pz, CPz) were chosen within 300 to 500 ms for P300.

## 3. Results

### 3.1. Measures of Acute Subjective Effects

We investigated the effect of the drug conditions (DMT/HAR, HAR, PLA) on the elementary and complex imagery scores using rmANOVA. There was a statistically significant effect of drug condition on elementary imagery scores (*F*(1.23, 36.891) = 42.314, *p* <0.001, *η*^2^*p* = 0.585). The post-hoc test revealed that DMT/HAR significantly increased the elementary imagery scores (Mean= 43.2, SD=33) compared to PLA (*p*< 0.001; Mean= 0.12, SD=0.7) and HAR (*p*< 0.001, Mean= 4.2, SD=13.2) (Figure 1). There was also a statistically significant effect of drug condition on complex imagery scores (*F*(1.371, 41.141) = 47.524, *p*< 0.001, *η*^2^*p* = 0.613). The post-hoc tests revealed that DMT/HAR significantly increased the complex imagery scores (Mean= 51.1, SD=34.2) compared to PLA (*p*< 0.001; Mean= 0.39, SD=1.9) and HAR (*p*< 0.001, Mean= 6.9, SD=18.3; Figure 1). There were no significant differences between the PLA and HAR conditions on either elementary or complex imagery scores (all *p* > 0.05).

**Figure 1.**
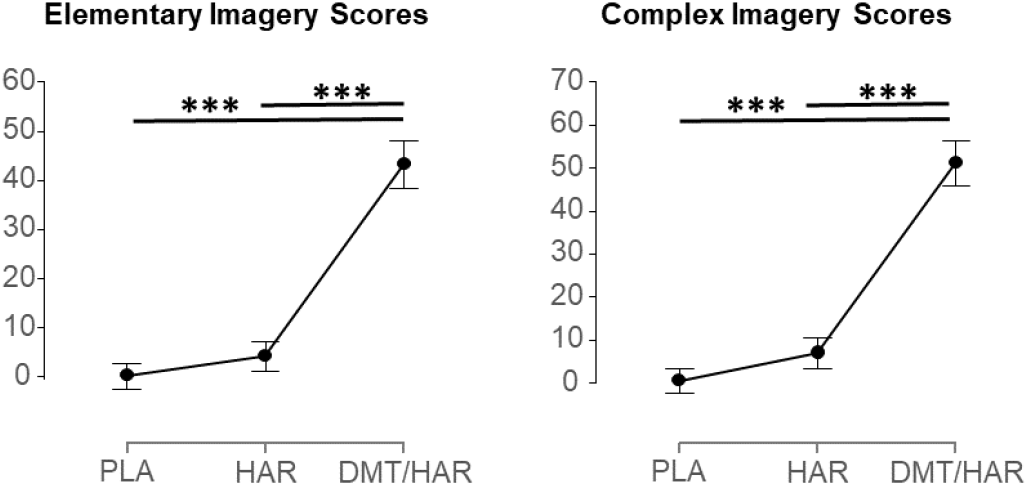
Plots showing the elementary and complex imagery scores. DMT/HAR significantly increased elementary and complex imagery scores (DMT/HAR vs. PLA and DMT/HAR vs. HAR: p<0.001; PLA vs. HAR: n.s.).

### 3.2. Face Processing EEG experiment

#### Self Face

First, we investigated the neural response to self-faces. A nonparametric cluster-based permutation analysis was run to investigate the effect of drug conditions (DMT/HAR, HAR, and PLA) on the ERPs in response to self-face category (refer to Table S1 in *Supplementary Materials* for all statistical results). Cluster-based analysis (MANOVA, comparing the three drug conditions) revealed 2 positive clusters at 140–199 ms and between 207–600 ms (all p<0.001), suggesting a statistical difference between the responses to self-faces across drug conditions. The direct comparison revealed that the DMT/HAR condition increased the neural response to self-face within the time window of 140–200 ms at posterior-occipital electrodes compared to both PLA and HAR conditions (DMT/HAR vs. PLA, p=0.022; DMT/HAR vs. HAR, p<0.001; Figure 2 A, B). The same analysis revealed a reduction of N170-P300 waves in the DMT/HAR condition compared to the other two drug conditions (Figure 2 B). In addition, DMT/HAR increased neural responses within the time window of 200–600 ms at the frontal electrodes compared to the other drug conditions (DMT/HAR vs. PLA, all clusters p<0.025; DMT/HAR vs. HAR, p<0.001; see Figure 2 A). There was no significant difference between HAR and PLA.

**Figure 2.**
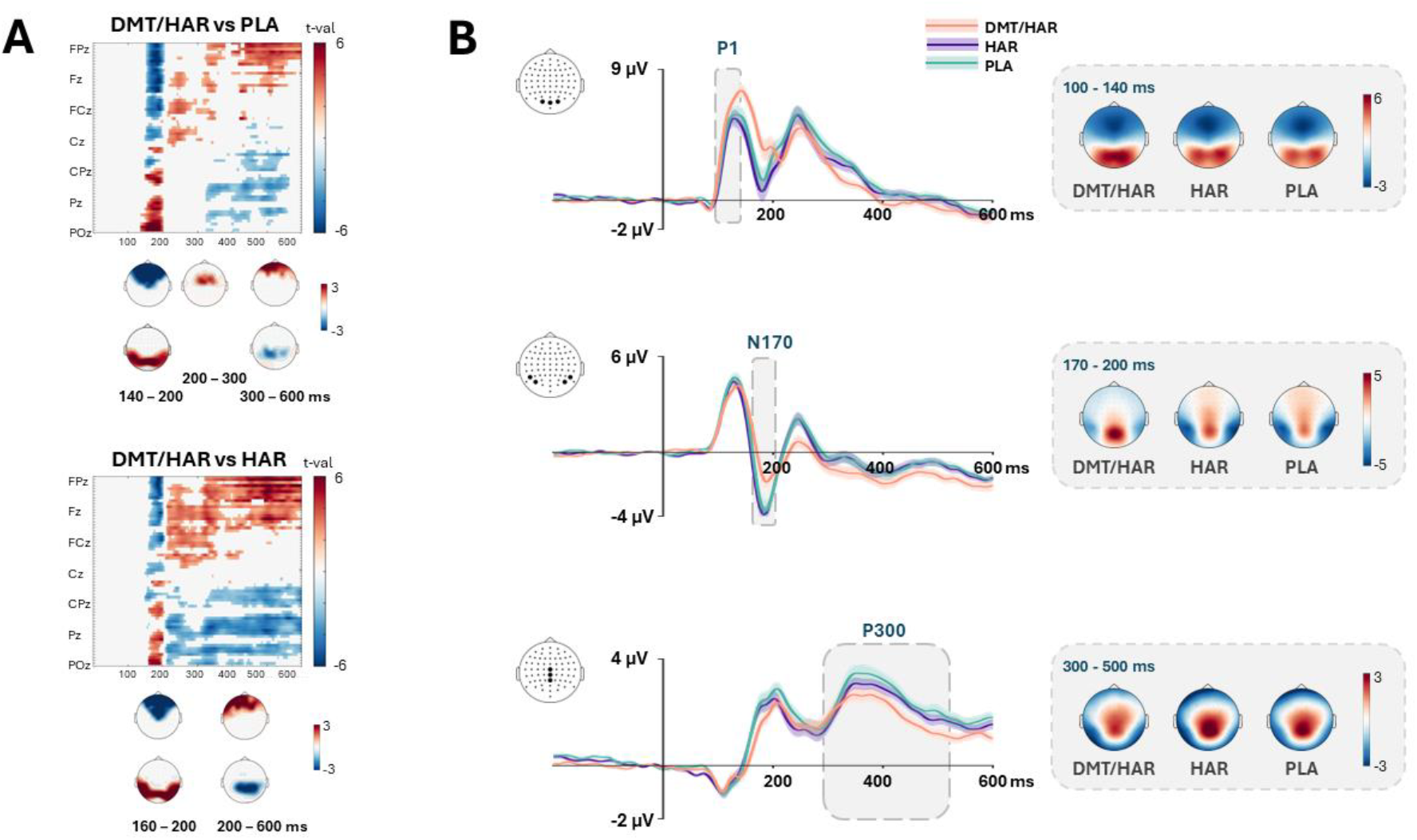
The effect of DMT/HAR on self-face. Spatiotemporal matrices display the results of the cluster-based permutation test, comparing the responses across conditions (DMT/HAR vs. PLA and DMT/HAR vs. HAR) to self-face stimuli. Significant clusters are shown as topographic distributions at the bottom. (B) Grand averaged ERPs (mean and SE) computed across all participants, averaged over O1, O2, Oz for P1 (upper), PO8, P8, PO7, P7 for N170 (middle) and Pz, CPz, Cz for P300 (lower) and scalp topography of each drug condition averaged within relevant time-windows on the right side. Electrodes were chosen based on the results of the cluster-based permutation test. DMT/HAR decreased N170 and P300 compared to both HAR and PLA. No significant differences emerged between HAR and PLA.

#### Familiar Face

The same analytical approach was performed on the familiar face category. Cluster-based analysis (MANOVA) revealed 2 positive clusters between 80–400 ms and 400–580 ms (p<0.001 and p=0.017), suggesting a statistical difference between the response to familiar face images as a function of drug condition. The direct comparison revealed that DMT/HAR increased the neural response to familiar faces within the time window of 90–200 ms at the posterior-occipital electrodes compared to the other drug conditions (DMT/HAR vs. PLA, p=0.013; DMT/HAR vs. HAR, p=0.018) (Figure 3 A). The effect revealed altered P1-N170 waves in the DMT/HAR condition compared to both PLA and HAR conditions (Figure 3 B). In addition, DMT/HAR increased neural responses within the time window of 200–600 ms at frontal electrodes compared to HAR (all clusters p<0.025) but not when compared to PLA (p>0.025) (Figure 3 A). Interestingly, HAR increased the neural response to familiar faces within the time window of 265–480 ms compared to PLA (p=0.01) at right central-parietal-temporal electrodes (see Figure S1 in *Supplementary Materials*).

**Figure 3.**
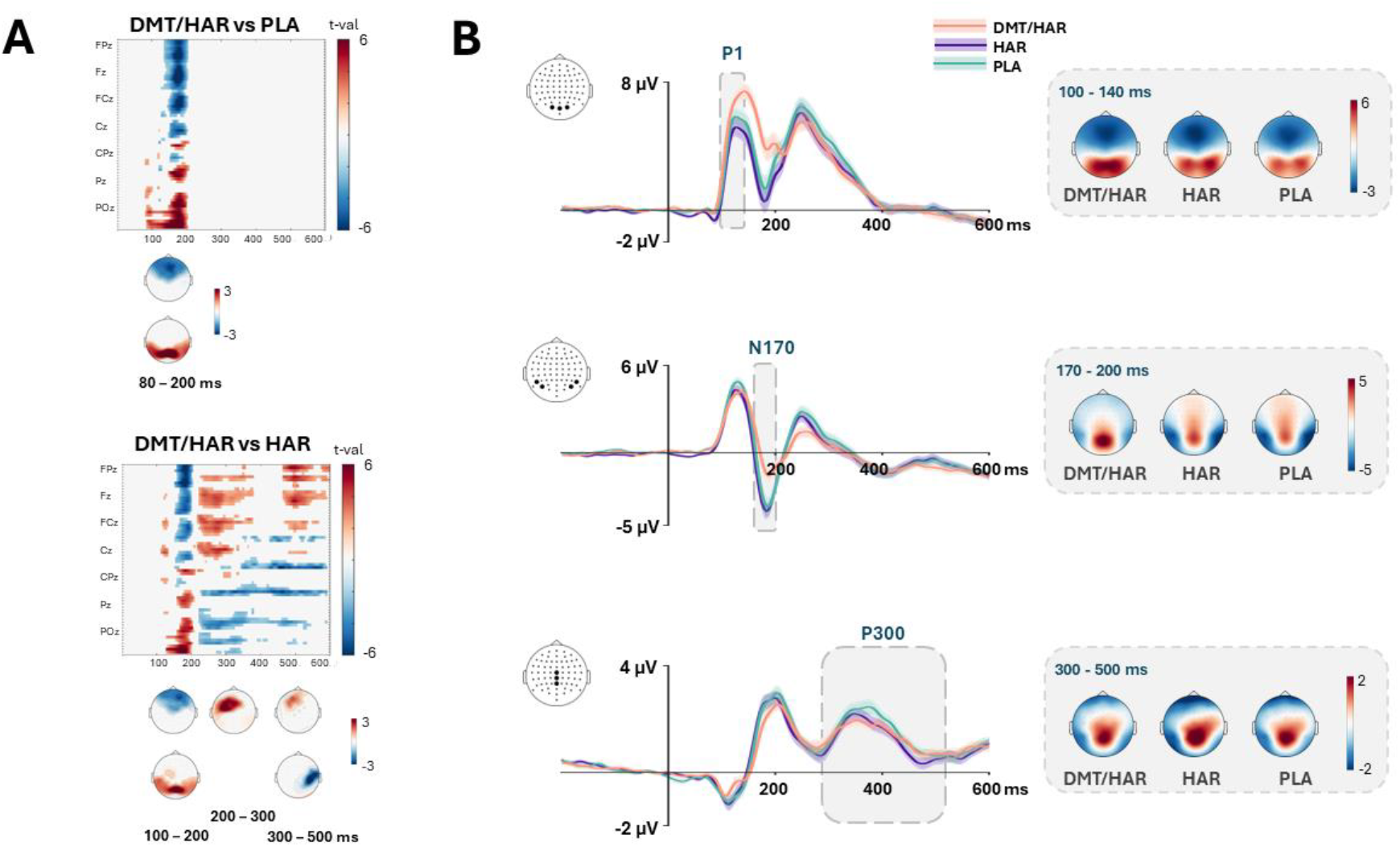
The effect of DMT/HAR on familiar faces. (A) Spatiotemporal matrices display the results of the cluster-based permutation test, comparing the response to familiar face stimuli across drug conditions (DMT/HAR vs. PLA and DMT/HAR vs. HAR). Significant clusters are shown as topographic distributions at the bottom of the graphs. (B) Grand averaged ERPs (mean and SE) computed across all participants, averaged over O1, O2, Oz for P1 (upper), PO8, P8, PO7, P7 for N170 (middle) and Pz, CPz, Cz for P300 (lower) and scalp topography of each drug condition at relevant time windows on the right side. Electrodes were chosen based on the results of the cluster-based permutation test. DMT/HAR elicited increased P1 and decreased N170 compared to both HAR and PLA. DMT/HAR did not affect P300.

#### Unknown Face

The same analytical approach was performed on the unknown face category. Cluster-based analysis (MANOVA) revealed 2 positive clusters at 125–205 ms and between 215–600 ms (all p<0.001), suggesting a statistical difference in drug conditions on an unknown face. The direct comparison revealed that DMT/HAR increased the neural response to an unknown face within the time window of 130–205 ms at the posterior-occipital electrodes compared to the PLA condition (p= 0.02) but not when compared to HAR (p> 0.025) (Figure 4 A). The effect revealed altered N1-P300 waves in the DMT/HAR condition compared to the PLA condition (Figure 4 B). In addition, DMT/HAR decreases neural responses within the time window of 300–600 ms at the right frontal central posterior temporal electrodes compared to both PLA (p< 0.001) and HAR (p= 0.004).

**Figure 4.**
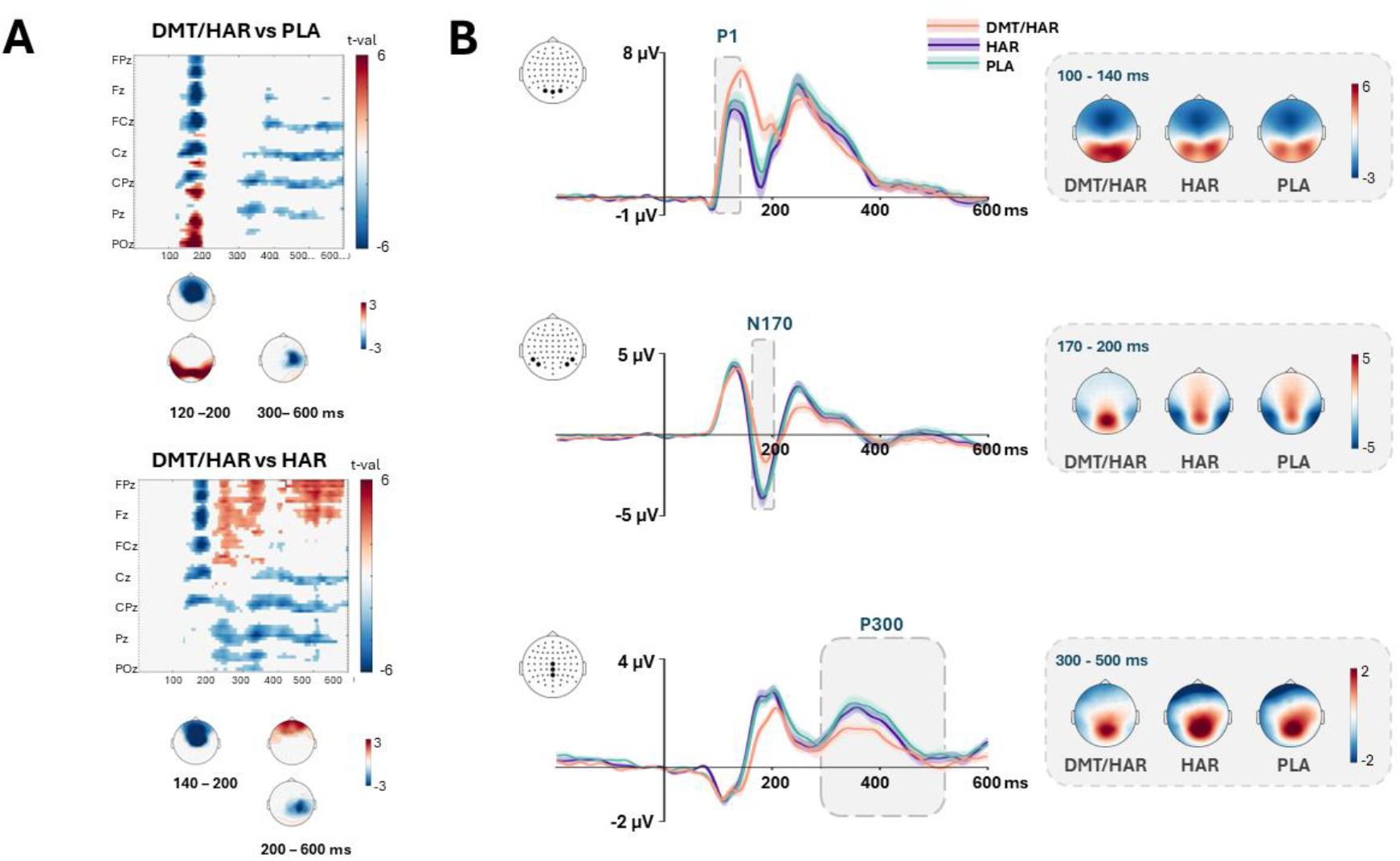
The effect of DMT/HAR on unknown faces. (A) Spatiotemporal matrices display the results of the cluster-based permutation test, comparing the conditions (DMT/HAR vs. PLA and DMT/HAR vs. HAR) response to unknown face stimuli. Significant clusters are shown as topographic distributions at the bottom of the graphs. (B) Grand averaged ERPs (mean and SE) computed across all participants, averaged over O1, O2, Oz for P1 (upper), PO8, P8, PO7, P7 for N170 (middle) and Pz, CPz, Cz for P300 (lower) and scalp topography of each drug condition at relevant time windows on the right side. Electrodes were chosen based on the results of the cluster-based permutation test. DMT/HAR elicited decreased N170 and P300 compared to PLA and decreased P300 compared to HAR. There were no significant differences between PLA and HAR drug conditions.

#### Interim Summary

In summary, our investigation revealed a similar impact of DMT/HAR on ERP components within the first 200 milliseconds across all face categories (self, familiar, and unknown). During this time, there was a selective DMT/HAR-induced increase in neural response at parieto-occipital electrode sites compared to both placebo (PLA) and harmine (HAR). In the next section, we investigated whether the influence of psychedelics distinctly modifies the processing of one’s own face in comparison to familiar and unknown faces.

#### ‘Does DMT/HAR specifically modify self-referential information processing?’

We hypothesized that DMT/HAR would attenuate the processing of self-relevant facial stimuli compared to other facial categories, thereby reducing brain differentiation between self and other faces. A nonparametric cluster-based permutation analysis was run for the interaction effect between drug conditions and the differential ERP waves calculated by subtracting the signal between face categories (e.g., *DMT/HAR*_*self−unknown*_ vs. *PLA*_*self−unknown*_; *DMT/HAR*_*self−unknown*_ vs. *HAR*_*self−unknown*_; *etc*.) (refer to Table S2 in *Supplementary Materials* for all the detailed statistical results).

The results confirmed the hypothesis, showing that the differential EEG responses to self-minus-unknown and self-minus-familiar faces were significantly reduced in DMT/HAR compared to both PLA and HAR. This corresponded to a negative cluster approximately between 320 and 550 ms (Self-Unknown: DMT/HAR vs. PLA, p=0.024; Self-Familiar: DMT/HAR vs. PLA, p= 0.004; DMT/HAR vs. HAR, p= 0.002) comprising several central-posterior electrodes (Figure 5 A). These results suggest that the neural response measured at centro-posterior electrodes when processing self-faces was more similar to familiar and unknown faces under the effect of DMT/HAR. A post-hoc cluster-based analysis was run to compare amplitudes of responses to self, familiar and unknown faces within a 300–500 ms time window (comprising the P300) at the central posterior electrodes (CPz, POz, Pz). Results revealed that the neural response to self-faces was greater in PLA and HAR compared to unknown and familiar faces. This increase in neural response for self faces was substantially reduced in DMT/HAR(Figure 5 C).

**Figure 5.**
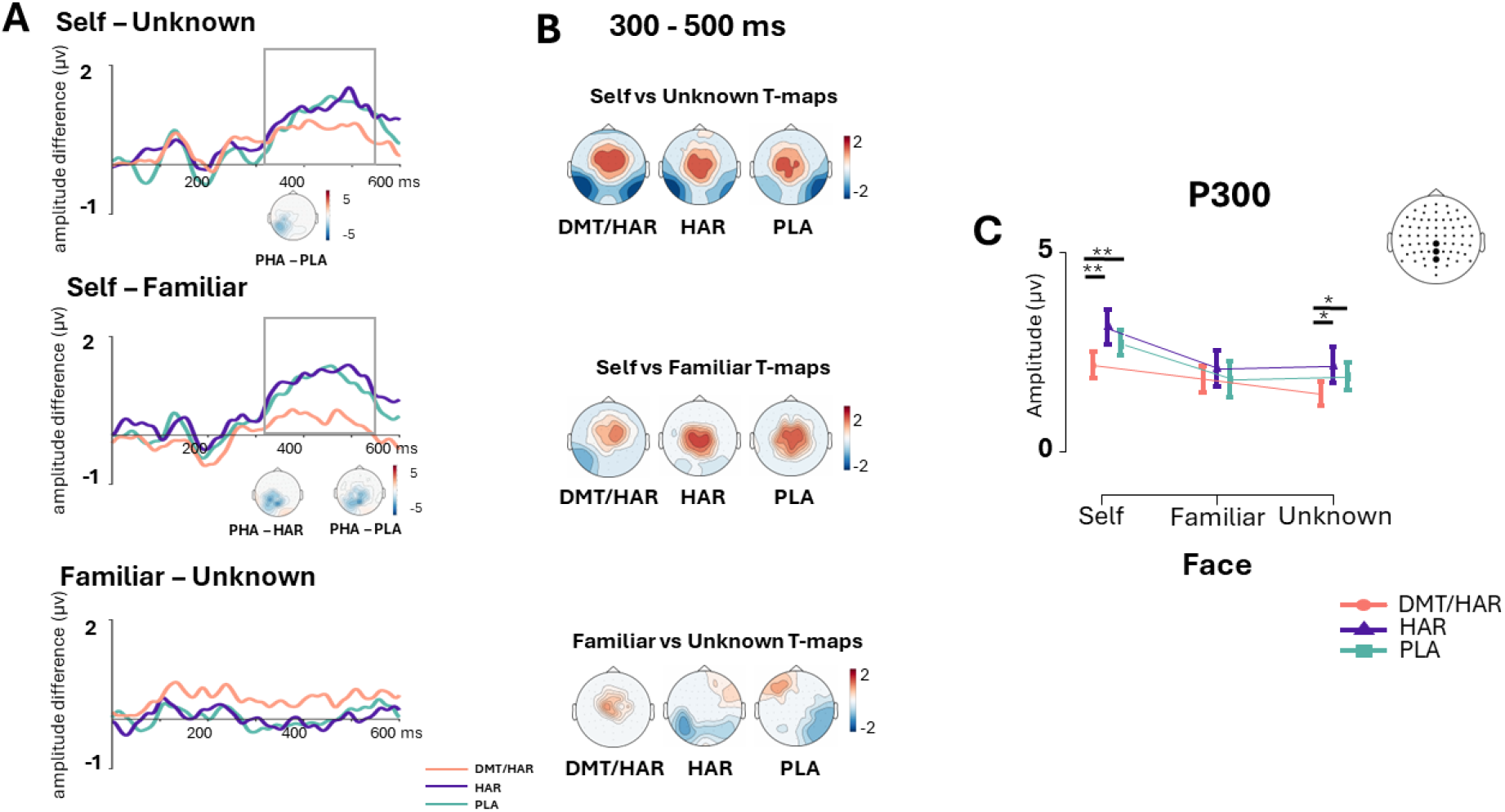
Interaction effect. Plots showing the significant differential ERPs of Self-Unknown (upper) and Self-Familiar (middle), and Familiar-Unknown (lower) at the posterior electrodes (POz, Pz, CPz) within 300–550 ms. Topographies of significant interaction effects of drug conditions are shown below the differential ERP plots. (B) Topographies of the difference responses for Self - Unknown, Self - Familiar and Familiar – Unknown for each drug condition (DMT/HAR, HAR, PLA). (C) Interaction plots show decreased responses to self and unknown faces in DMT/HAR condition compared to HAR and PLA conditions during the P300 time window over central-posterior electrodes (POz, Pz, CPz). Results revealed that the DMT/HAR condition attenuated the neural response to self-faces, and in turn, the differences between neural responses to self-faces and the other face categories were reduced.

#### Correlations between EEG and Psychometric Data

Without any a priori hypothesis, a cluster-based correlation was performed between differential EEG data (DMT/HAR-PLA) and elementary/complex imagery scores (DMT/HAR-PLA) across all electrodes and all time points (0–600ms). No significant correlation-clusters were observed for the entire time window (0–600 ms; all p>0.05). Additionally, no correlations were observed between the elementary/complex imagery scores and enhanced P1 (90–140 ms)/decreased N170 (170–200 ms) and reduced P300 (300–500 ms) components (all p>0.05).

## 4. Discussion

The results of this placebo-controlled, double-blind, within-subject randomized design study provided novel insights into psychedelics’ effects on the neurophysiological signatures of face recognition. Our findings demonstrate significant alterations in early visual processing during the psychedelic experience induced by novel ayahuasca-inspired DMT/HAR formulation. Most importantly, we observed specific modulation of the self-face perception compared to familiar and unknown faces, emphasizing the effect of psychedelics on self-referential information processing.

### General Effects of Psychedelics on Early Visual Processing

Our study’s primary finding is that DMT/HAR administration enhances neural activity in the parieto-occipital cortex during the early stages of visual perception (<200 ms) for all face categories (Self, Familiar, Unknown). This time frame includes both the early P1 and N170 waves of event-related potentials (ERPs). Specifically, DMT/HAR increases the P1 amplitude at medial occipital electrodes while decreasing the N170 amplitude at lateral occipital-temporal electrodes. These changes in early visual ERP components – marked by an increased P1 and a reduced N170 – indicate that DMT/HAR influences face processing differently depending on the stage of visual analysis.

This finding shows that the effect of psychedelics can be dissected within the chronology of face perception, ultimately expanding our understanding of the differential effects of serotonergic modulation during visual processing. 5-HT2A receptors are highly expressed in the visual cortex (Dyck & Cynader, 1993; Gerstl et al., 2008; Watakabe et al., 2009). Within the ventral occipitotemporal face processing network, faces are processed across different selective regions. The information process initiates in the inferior occipital region (“occipital face area”, OFA), which is sensitive to the low-level features of the visual input (e.g., size, luminance, contrast, spatial frequency spectrum). The activity of this region has been associated with the P1 ERP component. Subsequently, the information reaches the lateral middle fusiform gyrus (commonly referred to as the fusiform face area, FFA) and the posterior superior temporal sulcus (pSTS), which are responsible for high-level structural encoding of faces and linked to the N170 component of the ERPs (Duchaine & Yovel, 2015; Haxby et al., 2000; Rossion & Jacques, 2011; Sadeh et al., 2010; Tsao & Livingstone, 2008). Our results indicate that DMT/HAR alters the core system of face processing by increasing the P1 component, resulting in an enhanced visual gain while impairing the structural encoding of faces as captured by the attenuated N170 component. Similar findings from previous studies (Kometer et al., 2011, 2013) using non-face stimuli suggest that the altered responses observed as increased or decreased early visual ERP components might not be exclusive to face processing. Instead, these changes could reflect a broader effect of psychedelics on early visual processing. For example, Kometer et al. (2011) demonstrated that psilocybin dose-dependently increased the medial P1 and decreased the N170 during an object completion task. Furthermore, research utilizing a forced-choice direction of motion discrimination task revealed that psilocybin specifically impairs coherence sensitivity for random dot patterns, likely affecting high-level global motion detectors. In contrast, there was no notable impact on contrast sensitivity for drifting gratings, which are typically governed by low-level detectors (Carter et al., 2004).

Integrating these EEG findings – where psychedelics increase the P1 component and decrease the N170 – suggests that psychedelics may differentially influence low-level and high-level visual processing. Specifically, they seem to enhance the perception of basic visual features (e.g., brightness, contrast, local motion detection) while disrupting higher-level integration processes (e.g., structural encoding of faces, object recognition, global motion detection). These observations provide a compelling rationale for understanding the emergence of elementary visual experiences under psychedelics. In contrast to previous studies that linked increased medial P1 to enhanced brightness perception and decreased N170 to visual hallucinations (Kometer et al., 2011b, 2013b), our study did not find a statistically significant correlation between ERP component changes and scores related to complex and elementary imagery. This was despite higher subjective ratings of visual distortions in the DMT/HAR condition. Therefore, further investigation is warranted to elucidate the neural mechanisms underlying elementary imagery and to explore the neural basis of complex imagery – including the visualization of autobiographical memories or unfamiliar entities and locations – which remains a complete mystery (Davis et al., 2020; Lutkajtis, 2021).

### Specific effects of Psychedelics on Self-referential Information Processing

Next, we examined the specific modulation of self-referential information processing under psychedelics. The phenomenology of psychedelics is marked by profound changes in self-experience, including the dissolution of the self or ego and the blurring of boundaries between the self and the external world. These experiences often foster a heightened sense of empathy and compassion towards oneself and others (Blatchford et al., 2020; Letheby & Gerrans, 2017; Tagliazucchi et al., 2022). According to social neuroscience, prosocial traits such as empathy and compassion are rooted in shared representations of the self and others (Preston & Hofelich, 2012). Based on these insights, we hypothesized that the DMT/HAR condition would show similar ERP responses to self-faces as to familiar or other-related faces, resulting in smaller ERP differences between these face categories. Consistently, the spatiotemporal analysis of EEG signals across the scalp and throughout the time window (0–600 ms) revealed a significant drug-by-face interaction. The DMT/HAR condition notably diminished the neural response difference between self and unknown or familiar faces at posterior midline electrodes during the 300–500 ms timeframe, corresponding to the P300 ERP component. This effect was observed when compared to both harmine and placebo conditions (Figure 5).

The P300 ERP component, known for reflecting attentional resource allocation (Polich, 2007, 2012), is typically associated with activations in the default mode network (DMN) structures related to self-referential processing (Knyazev, 2013). Self-relevant information, due to its high social and adaptive value, generally receives prioritized access to attentional resources (Gray et al., 2004). Electrophysiological studies consistently show that the P300 amplitude is enhanced in response to self-referential stimuli, such as one’s own face, compared to famous or unknown faces (Scott et al., 2005; Sui et al., 2006; Tacikowski & Nowicka, 2010), as well as to one’s own name versus other names (Tacikowski et al., 2011, 2014), even during sleep (Perrin et al., 1999) and unconscious perception (Doradzińska et al., 2020). Our results indicate that DMT/HAR markedly diminished the P300 amplitude for self-faces, and to a lesser extent for unknown faces, relative to both harmine and placebo. Notably, the P300 amplitude for familiar faces remained stable across all drug conditions (Figure 5C). These findings suggest a significant reduction in attentional focus on self-relevant information during the psychedelic experience, which resulted in reduced differentiation between self-other and self-familiar faces.

Previous research has shown similar effects of psychedelics on self-referential information processing. For example, a recent EEG-ERP study found that psilocybin reduced the difference in P300 responses between self and other stimuli during an auditory self-monitoring task (Smigielski et al., 2020). This reduction in P300 amplitude was significantly correlated with the intensity of experiences related to unity and altered perception. Interestingly, similar reductions in P300 amplitude were observed in individuals practicing Loving-Kindness Meditation (LKM), where smaller differences in P300 responses between self and other stimuli were inversely related to the extent of meditation experience (Trautwein et al., 2016). These findings suggest that both psychedelics and meditation can influence self-referential processing and perception in comparable ways, potentially through shared mechanisms affecting self-awareness and cognitive integration (Holas et al., 2023; Millière et al. 2018). Specifically, both approaches appear to diminish the distinction between self and familiar or unknown stimuli during the P300 processing stage. This alteration in neural processing might be linked to the heightened feelings of unity, compassion, and empathy often reported during psychedelic experiences (Preller & Vollenweider, 2018).

The current study provides evidence that psychedelics reduce the focus on the self and lead to a stronger self–other integration, which has been thought to be an underlying mechanism of improved social and emotional cognition (Dinulescu et al., 2021). Such a mechanism would have important implications as changes in self-processing might play a vital role in the effectiveness of psychedelic-assisted therapy (Eisner & Cohen, 1958; Nayak & Johnson, 2021; Reiff et al., 2020). Many mental disorders, including depression and anxiety, are often characterized by excessive negative-biased self-focus (rumination), feelings of isolation, and decreased attention to others and the environment, which is attributed to heightened DMN resting-state activity and an altered balance between DMN and executive network activity (Brakowski et al., 2017; Coutinho et al., 2016; Dixon et al., 2022; Palhano-Fontes et al., 2015). The temporary dissolution of self through psychedelics may lead to more flexible cognitive responses, particularly for individuals who experience negative self-attribution and ruminative thinking (Fauvel et al., 2023). Accordingly, it has been shown that psilocybin significantly decreases depression and anxiety in life-threatening cancer patients (Griffiths et al., 2016; Ross et al., 2016). Moreover, the encounter with self-dissolution, a sense of oneness, and disembodiment culminates in a realization that the intricate texture of emotions and thoughts is, in fact, nothing but a construction. This transformative shift in perception might serve as a catalyst for a more profound sense of self and existence, effectively dismantling dysfunctional self-perceptions. Furthermore, it might promote greater empathy toward others and facilitate social adaptation (Dolder et al., 2016; Pokorny et al., 2017), considered a key to authentic and enduring happiness (Dambrun & Ricard, 2011).

### Limitations and Future Directions

While our study has provided significant insights into the effects of psychedelics on face recognition, it is crucial to recognize that the observed alterations in these processes may be influenced by the specific characteristics of the novel DMT/HAR formulation and the dosage utilized in our study. Variations in psychedelics or dosages may result in diverse effects on face processing. Consequently, future research should focus on investigating the generalizability of our findings to different psychedelics and doses, as well as the effects at different timepoints, aiming to gain a comprehensive understanding of the broader impact of psychedelics on face perception and recognition.

Secondly, we did not find any correlations between subjective effects (elementary and complex imagery scores) and electrophysiological data in this study. One possibility is that variations in how participants individually perceive psychedelics, including differences in the type and intensity of visual hallucinations experienced, may play a significant role in explaining the observed absence of correlation. The inherently subjective nature of the psychedelic experience, influenced by factors such as set and setting, may result in diverse perceptual outcomes among participants (Hartogsohn, 2017, 2022; Noorani, 2021). These variations may not align neatly with the chosen metrics, further complicating efforts to establish clear associations between subjective and neural responses. Future studies could benefit from employing more refined or additional measures to capture the nuanced interplay between subjective experiences and electrophysiological data.

Finally, in this study, celebrity faces were used as familiar stimuli. Given the expanding nature of the self (Aron & Aron, 2006; Aron & Fraley, 1999; Mattan et al., 2016), it has been proposed that personally familiar faces (e.g., a partner’s face, family member) may be processed not only as familiar faces but also as self-related stimuli, as if they were part of the self (Taylor et al., 2009). It would be interesting to explore the potential overlap between personally familiar faces and self-related stimuli during psychedelic experiences to better understand the intricate relationship between psychedelics, face recognition, and self-referential processing.

## Conclusion

In conclusion, this pioneering study provides valuable insights into the effects of psychedelics on self-face perception and recognition. Using a data-driven approach and taking advantage of the high temporal resolution of EEG, we observed the effect of an ayahuasca-inspired DMT/HAR formulation on early visual processing, namely increased P1 and attenuated N170 across all face categories. Furthermore, we discovered that DMT/HAR specifically modulated self-referential processing by reducing neural activation in response to the self-face, as revealed by the alteration of the P300 wave. These findings underscore the modulatory role of serotonergic neurotransmission in visual processing and face recognition. Our results make a significant contribution to the existing knowledge about how serotonergic psychedelics affect visual and self-face processing, offering potential implications for disorders characterized by disturbances in self-referential processing. Further research in this area holds promise for advancing our understanding and exploring potential therapeutic interventions.

## Supporting information

Supplementary Meterials

## Data Availability

All data produced in the present study are available upon reasonable request to the authors

## Author contributions

DS_1,2,3,4,5,6,7,8_, HDA_1,2,3,7,8_, MK_1,5,8_, MJM_8_, LC_3,8_, AH_3,8_, CPS_3,8_, CE_3,8_, IAW_3,8_, JM_3,8_, DM_3,8_, DAD_1,8,9_, MS_1,2,7,8_, DB_1,5,7,8_^1^ conception and design of the study; ^2^ study management; ^3^ data acquisition; ^4^ data analysis, ^5^interpretation of results; ^6^ drafting the article; ^7^ revision of the article; ^8^ final approval of the version to be submitted; ^9^ Manufacturing of study drug

## Acknowledgements

We extend our gratitude to Hanspeter Landolt for providing the research facility and infrastructure. We also wish to thank our study participants for their valuable contributions to this research.

## Research ethics and patient consent

The study was conducted according to the World Medical Association Declaration of Helsinki.

