## Supplementary Meterials for "Ayahuasca-Inspired DMT/HAR Formulation Reduces Brain Differentiation Between Self and Other Faces"

**Supplementary Materials**

**Table S1-** Results of cluster-based permutation tests on the event-related potentials showing the significant effect of drug condition (DMT/HAR, HAR, PLA) on self, familiar and unknown faces.

| **Main Effect** | **Comparisons** | **Probability** | **Statistics** | **Time (ms)** |
| --- | --- | --- | --- | --- |
| *Self-Face* | MANOVA | p<0.001 p<0.001 | Tsum=41383.9 Tsum=13633.7 | 140 - 199 ms 207 - 600 ms |
|  | DMT/HAR vs. PLA | p=0.002997 p=0.015984 p=0.021978 p=0.004995 p=0.008991 | Tsum=1852.0 Tsum=1187.2 Tsum=1046.8 Tsum=-2041.2 Tsum=-1662.2 | 398 - 601 ms 214 - 378 ms 136 - 199 ms 324 - 562 ms 148 - 203 ms |
|  | DMT/HAR vs. HAR | p<0.001 p<0.001 | Tsum=6559.2 Tsum=6278.1 | 160 - 601 ms 148 - 601 ms |
|  | HAR vs. PLA | n.s. |  |  |
| *Familiar Face* | MANOVA | p<0.001 p= 0.017 | Tsum=31602.2 Tsum= 4250 | 82 - 398 ms 406 - 582 ms |
|  | DMT/HAR vs. PLA | p=0.013 p=0.006 | Tsum=1572.8 Tsum= - 2080.2 | 82 - 207 ms 121 - 203 ms |
|  | DMT/HAR vs. HAR | p= 0.008 p= 0.017 p= 0.018 p= 0.001 p= 0.012 | Tsum=1719.6 Tsum=1117.1 Tsum=1070.4 Tsum= - 2662.5 Tsum= - 1290.3 | 218 - 378 ms 464 - 597ms 113 - 210 ms 222 - 589 ms 152 - 203 ms |
|  | HAR vs. PLA | p= 0.01 | Tsum=1115.4 | 265 - 484 ms |
| *Unknown Face* | MANOVA | p<0.001 p<0.001 | Tsum=24755.2 Tsum=18174.4 | 214 - 601 ms 125 - 207 ms |
|  | DMT/HAR vs. PLA | p=0.02 p<0.001 p=0.003 | Tsum=1078.9 Tsum= - 2574.6 Tsum= - 2161.5 | 132 - 203 ms 296 - 601 ms 121 - 207 ms |
|  | DMT/HAR vs. HAR | p=0.004 p=0.004 p<0.001 | Tsum=1741.5 Tsum=1610.8 Tsum= - 5501.8 | 214 - 363 ms 398 - 585 ms 132 - 601 ms |
|  | HAR vs. PLA | n.s |  |  |

**Figure S1- The effect of harmine on familiar faces**

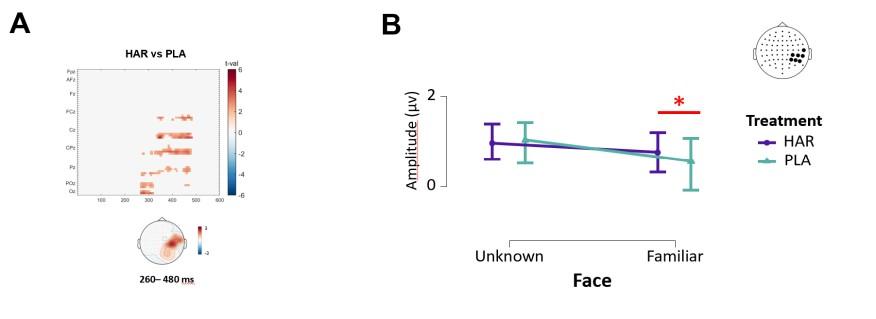
(A) Spatiotemporal matrix displays the result of the cluster-based permutation test, comparing the conditions HAR vs. PLA in response to familiar face stimuli. Significant clusters are shown as topographic distributions at the bottom of the matrix. (B) Interaction plots show that HAR increases the neural response to familiar faces (*p*=.004, 332-523 ms) at the right posterior temporal cortex (T8, TP8, CP6, CP4, P4, P6, P8) and while the response to unknown faces does not change when compared to the placebo condition.

**Table S2.** Results of cluster-based permutation tests on event-related potentials for interaction effects (one-tail = -1) testing the hypothesis of reduced differential brain responses to self vs. familiar faces and self vs. unknown faces.

| **Interactions** | **Comparisons** | **Probability** | **Statistics** | **Time (ms)** |
| --- | --- | --- | --- | --- |
| *Self vs. Unknown*  *Face*  *Self vs. Familiar Face*  *Familiar vs.*  *Unknown Face* | DMT/HAR vs. PLA | p=0.023976 | *Tsum*= -1267.5 | 312 - 562 ms |
|  | DMT/HAR vs. HAR | n.s. |  |  |
|  | HAR vs. PLA | n.s. |  |  |
|  | DMT/HAR vs. PLA | p=0.00399 | *Tsum*=-2035.2 | 324 – 554 ms |
|  | DMT/HAR vs. HAR | p=0.001998 | *Tsum*=-2705.9 | 317 - 600 ms |
|  | HAR vs. PLA | n.s. |  |  |
|  | DMT/HAR vs. PLA | n.s. |  |  |
|  | DMT/HAR vs. HAR | n.s. |  |  |
|  | HAR vs. PLA | n.s. |  |  |
